# 50 policies, 1 pandemic, 500,000 deaths: Associations between state-level COVID-19 testing recommendations, tests *per capita*, undercounted deaths, vaccination policies, and doses *per capita* in the United States

**DOI:** 10.1101/2020.09.04.20188326

**Authors:** Stephanie R. Perniciaro, Daniel M. Weinberger

## Abstract

**Background:** State health departments have been responsible for prioritizing and allocating SARS-CoV-2 tests and vaccines. Testing and vaccination recommendations in the United States varied by state and over time, as did vaccine rollouts, COVID-19 cases, and estimates of excess mortality.

**Methods:** We compiled data about COVID-19 testing, cases, and deaths, and excess pneumonia + influenza + COVID-19 deaths to assess relationships between testing recommendations, *per capita* tests performed, epidemic intensity, and excess mortality during the early months of the COVID-19 pandemic in the United States. We compiled further data about state-level SARS-CoV-2 vaccination policies and doses administered during the early months of the vaccine rollout.

**Results:** As of July 2020, 16 states recommended testing asymptomatic members of the general public. The rate of COVID-19 tests reported in each state correlated with more inclusive testing recommendations and with higher epidemic intensity. Higher *per capita* testing was associated with more complete reporting of COVID-19 deaths, which is a fundamental requirement for analyzing the pandemic. Testing *per capita* during the first three months was associated with vaccination *per capita* in the first three months of rollout. *Per capita* vaccine doses in each state were not associated with adherence to national guidelines.

**Conclusions:** Reported deaths due to COVID-19 likely represent an undercount of the true burden of the pandemic. States that struggled with testing rollout have also frequently struggled with vaccine rollout. Coordinated, consistent guidelines for COVID-19 testing and vaccine administration should be a high priority for state and national health systems.

## Introduction

SARS-CoV-2, the virus that causes COVID-19, was declared a global pandemic by the World Health Organization in March 2020.^1^ Worldwide, responses to the pandemic have varied and had varying success. In the United States, each state determined testing guidelines, lockdown procedures, and reopening procedures individually.^2^ The Centers for Disease Control and Prevention also maintained testing recommendations and guidelines, and produced guidance for establishing prioritized groups during vaccine rollout, but states and localities were largely responsible for their own policies.^3^ The COVID-19 pandemic in the United States is ongoing, costing over half a million lives by the end of February 2021. ^4^

Testing for COVID-19 has been a continual hurdle in the United States.^5^ Supply chain issues,^6^ doubts about testing accuracy and methodology,^7^ and conflicting guidance from federal, state and local officials about testing have all interfered with testing efforts.^8^ State governments and health departments established testing guidelines in March 2020, which were sometimes augmented by local governmental health departments.^9^ The turnaround time for test results was strained,^10^ and resources for individuals to sustain their household resources while remaining isolated or quarantined while awaiting the result of a test have not been widely available.^11^

This situation is now replaying with vaccine rollout and administration. Problems ranging from misinformation to convoluted registration systems have prevented eligible individuals from receiving vaccination appointments, echoed in problems including long wait times^12^ and sabotage^13^ hampering the actual administration of doses.

Here we describe the changing recommendations for COVID-19 testing to the general public in the United States in March-July 2020 and the vaccine recommendations in the December 2020-February 2021.

## Methods

The objectives of this analysis were to: 1) establish whether varying the state-level COVID-19 testing guidelines had any impact on the actual number of tests performed, and whether the number of tests performed was related to the rate of recognizing COVID-19 deaths as a cause of excess mortality (reported COVID-19 deaths/excess pneumonia + influenza+ COVID-19 (PIC) deaths) and 2) establish whether state-level vaccination policies had any impact on doses administered. COVID-19 testing guidelines were documented from the websites of the departments of health in each of the 50 states and the District of Columbia once monthly from March-July 2020 (Supplemental Table 1). We ranked recommendations to tests by the general public (as opposed to members of specific named risk groups) by assigning 0= criterion not specified, 1= accessible to people with a negative influenza test, 2= accessible to people with symptoms consistent with COVID-19 illness, 3= accessible to anyone, including asymptomatic people. Vaccination policies were recorded once monthly for December 2020, January and February of 2021.^14^ Adherence to Advisory Committee on Immunization Practices (ACIP) guidelines^15^ was scored for the first three phases of vaccine rollout as 0= more restrictive than ACIP guidelines/data not available, 1=identical to ACIP guidelines, 2=more inclusive than ACIP guidelines.

Correlations between testing recommendation score, tests *per capita*, and percentage of positive tests were analyzed with Spearman correlation tests, as were correlations between vaccine recommendation priority match with ACIP guidelines and number of doses administered total and *per capita*.

We evaluated relationships between the number of COVID-19 tests performed per capita, testing recommendations score, and epidemic intensity (defined as excess PIC deaths^16^per capita) using a negative binomial regression model. Total COVID-19 tests performed in each state was the outcome variable, offset by the log of the total population of each state, with the testing recommendations score (categorical) and the log of epidemic intensity as the independent variables.

We assessed the association of adherence to ACIP guidelines for vaccination prioritization and doses administered in a similar fashion, with doses administered as the outcome variable, offset by the log of the population, with adherence to ACIP guidelines and the current stage of vaccination rollout as the independent categorical variables. We investigated possible associations between testing *per capita* in March-May of 2020 and vaccine doses *per capita* in December 2020-February 2021 with monthly and overall Spearman correlation tests and with a negative binomial model as above where vaccine doses administered was the dependent variable, offset by the log of population, and the log of total tests administered/epidemic intensity was the independent variable.

We also used a generalized additive model (negative binomial link) to assess the completeness of death reporting. The outcome variable was the number of reported COVID-19 deaths in each state and month. The number of excess PIC deaths was included as an offset term, so this effectively models the ratio of reported COVID-19 deaths/excess PIC deaths. The *per capita* testing rate in each state and month was included as a penalized spline. Uncertainty in the estimates of excess PIC deaths were ignored in these analyses, and excess PIC deaths were constrained to be greater than or equal to the reported COVID-19 deaths. The model was fit using the package “mgcv” in R.

The number of excess PIC deaths per state and the reported number of COVID-19 deaths were obtained from a repository describing excess deaths during the COVID-19 outbreak (https://github.com/weinbergerlab/excess_pi_covid). ^16^ Positive tests and the overall number of tests in each state were collected from the website of The COVID Tracking Project (https://covidtracking.com/). Vaccine policy data were gathered from state health department websites, in a publicly available repository curated by the Kaiser Family Foundation.^17^ Vaccine uptake data were taken from the CDC’s COVID-19 Data Tracker (https://covid.cdc.gov/covid-data-tracker/#vaccinations) Population data were taken from the United States Census Bureau’s 2019 estimates. R (R: A language and environment for statistical computing, version 4.0.3 R Foundation for Statistical Computing, Vienna, Austria, https://www.R-project.org/.) was used for statistical analyses.

## Results

### State Testing Guidelines

From March to July 2020, recommendations of COVID-19 testing for the general public varied between states and over time (Figure 1), as did testing prioritization for identified risk groups (Supplemental Figure 1). As of July, 16 states recommended that asymptomatic members of the general public be tested for COVID-19. 9 states actively recommended against the testing of asymptomatic people; 3 of which then reversed this recommendation in July. South Dakota also recommended against seeking a test if people were experiencing symptoms, citing the lack of effective COVID-19 therapy as a contraindication for testing. Most states (35/51) changed their testing recommendations twice between March and July. Only Oklahoma did not change their testing criteria, though the state health department website specifies that private testing facilities have their own criteria.

**Figure 1.**
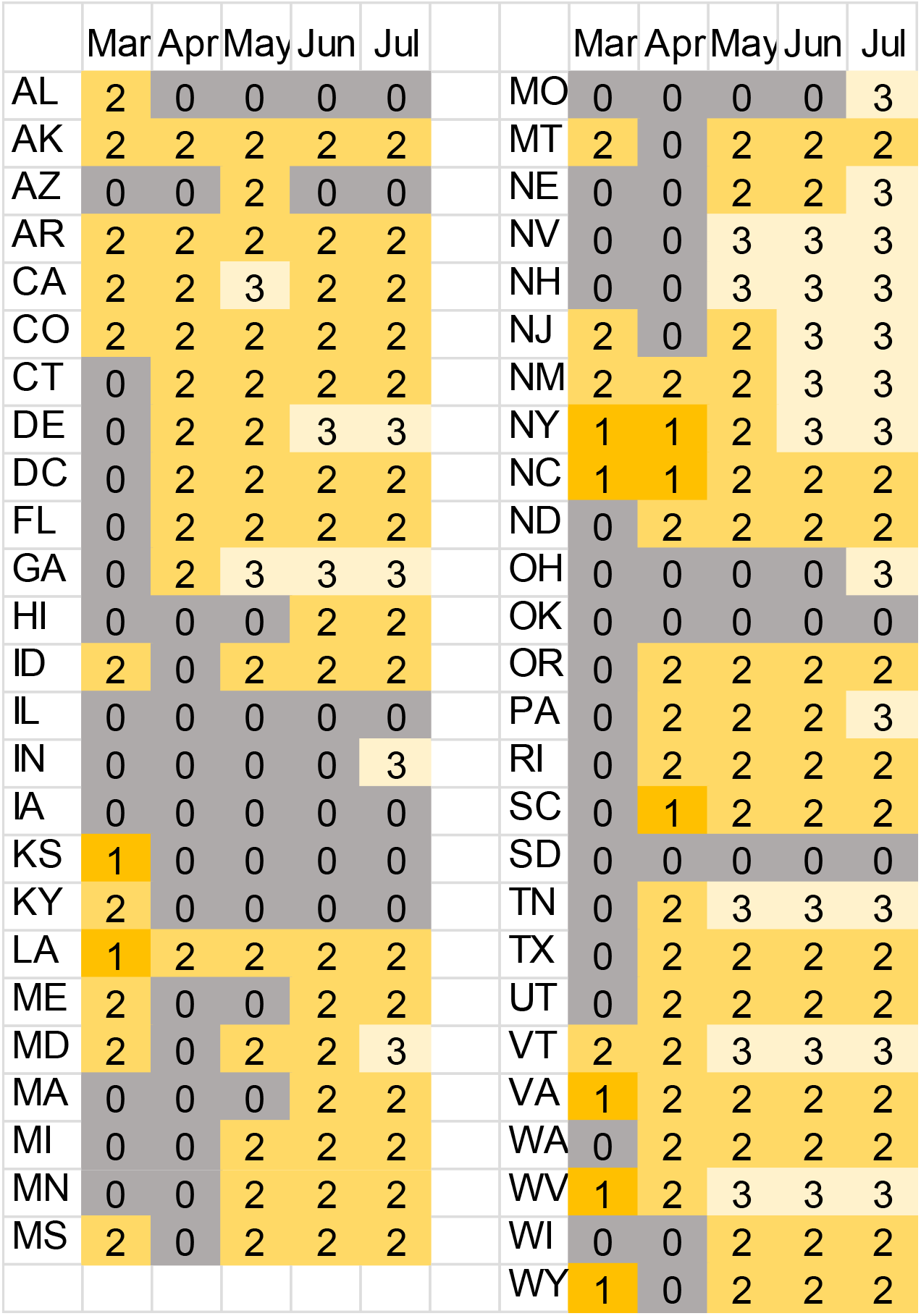
SARS-CoV-2 testing guidelines by state. Testing recommendations for the general public was scaled as follows: 0 = criterion not included, 1 = negative influenza test required, 2 = all symptomatic people, 3 = all people, including asymptomatic people. Testing guidelines for special risk groups are shown in Supplemental Figure 1.

### Factors associated with higher rates of testing

Reported COVID-19 tests *per capita* increased steadily, though unevenly, from March to June, while the percentage of positive tests varied widely between states (Figure 2, 3). States with more permissive testing recommendations had a lower percentage of positive tests (rho −0.22, 95% CI −0.34, −0.11), possibly reflecting the inclusion of lower risk individuals in the testing pool. The minimum percentage of positive tests for a single month was 0.8% in May in Alaska; the maximum was 47% in April in New Jersey. In a multivariate model, both more inclusive testing recommendations and higher epidemic intensity were associated with higher rates of testing.

**Figure 2.**
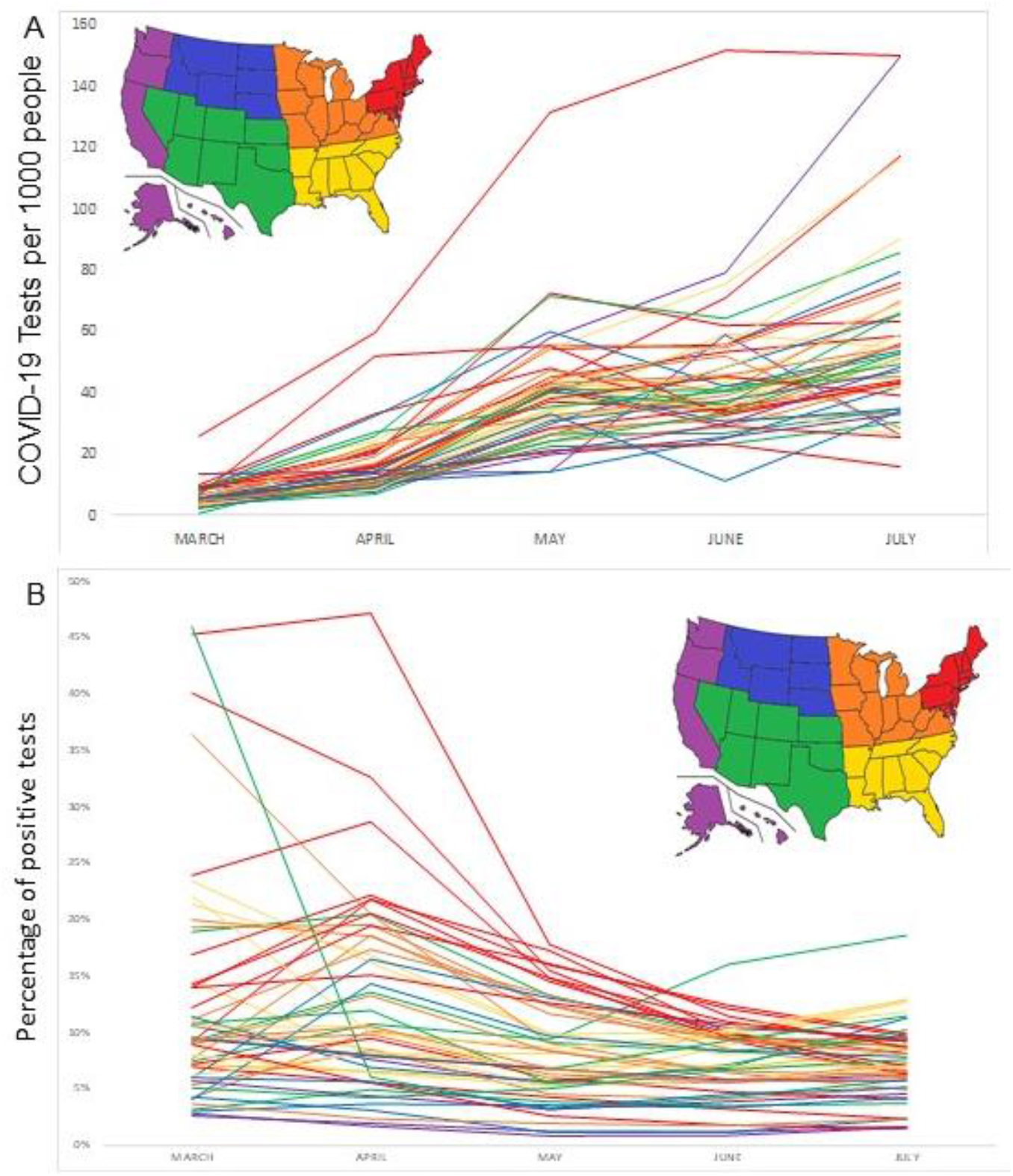
SARS-CoV-2 tests per 1000 population in the United States (A) and the percentage of positive SARS-CoV-2 tests (B), March-July 2020. Each state has a line; colors of lines are determined by geographic region per the inserted map.

**Figure 3.**
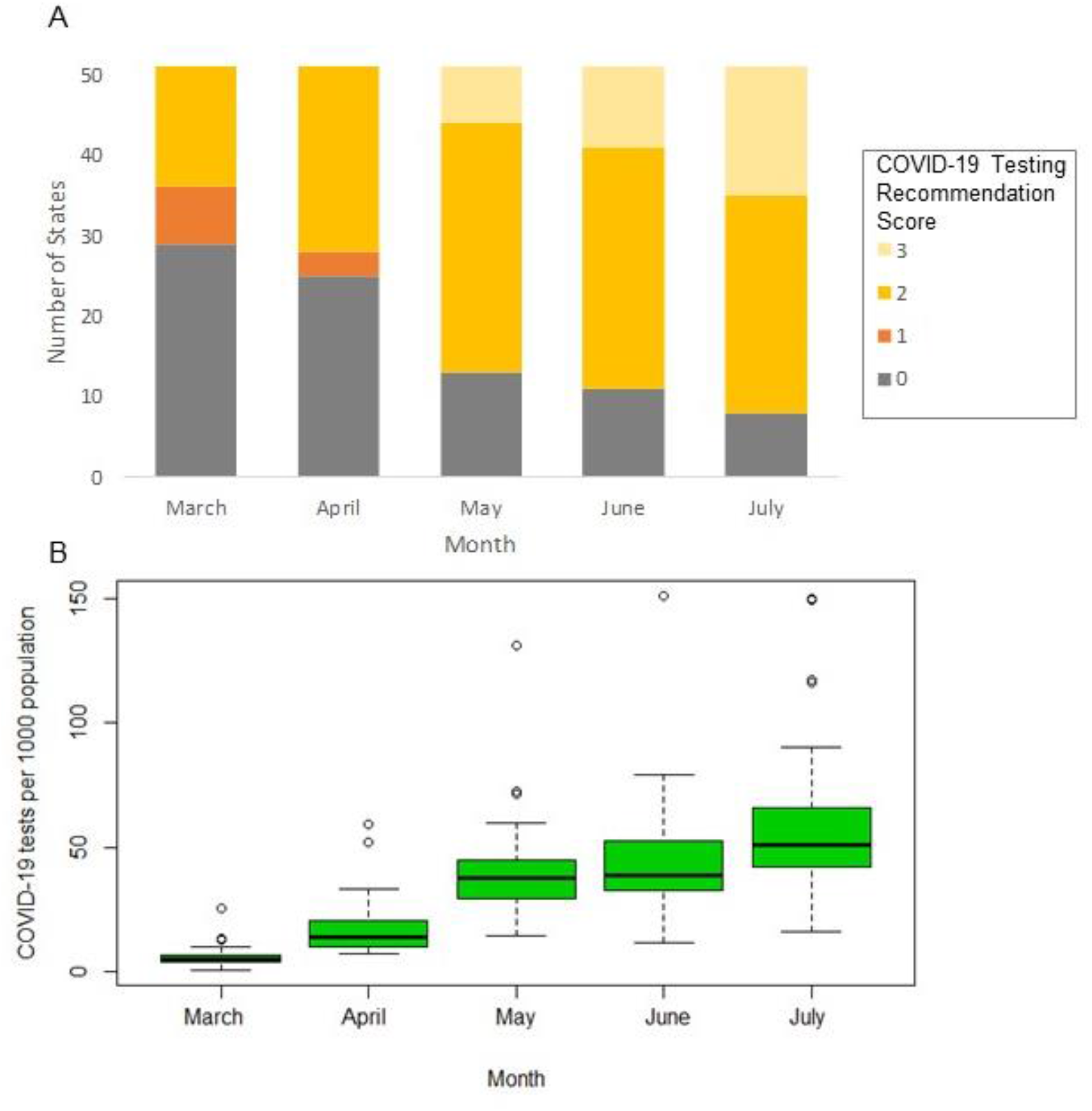
SARS-CoV-2 testing recommendation scores (A) and SARS-CoV-2 tests per 1000 people (B) from March-July, 2020 in the United States.

### Testing intensity and completeness of death reporting

The completeness of recording deaths as due to COVID-19 was related to the rate of testing. (Figure 4). This was particularly true in the first months of the pandemic when testing levels were low relative to the intensity of the epidemic; in later months, the level of testing was high and had little influence on the difference between reported COVID-19 deaths and excess PIC deaths.

**Figure 4.**
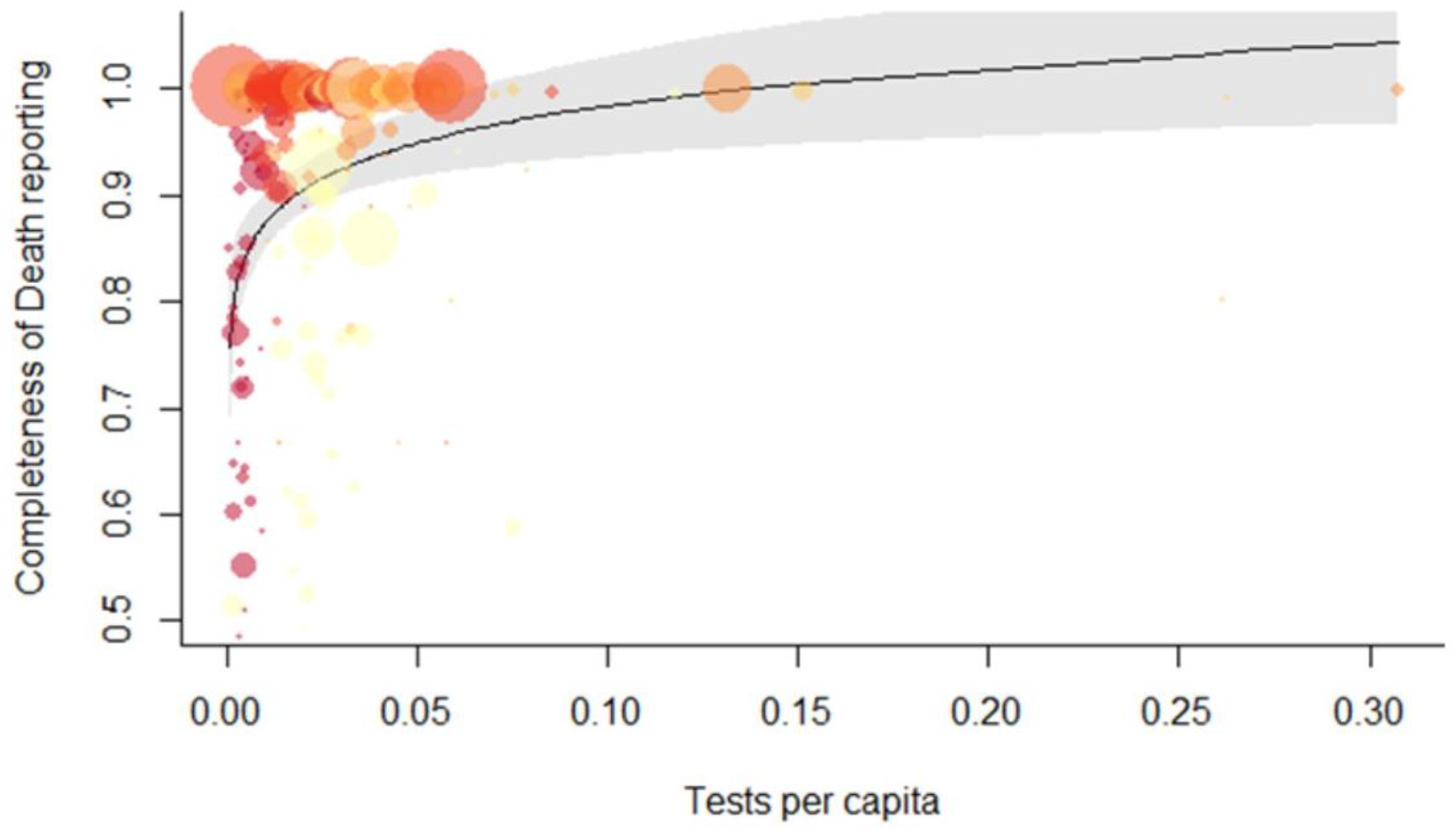
Association between the rate of testing for SARS-CoV-2 (per capita) and the completeness of death reporting, as measured by the ratio between reported COVID-19 deaths and excess deaths due to pneumonia/influenza/coronavirus (PIC). The line +/- 95% confidence intervals represents the trend as estimated from a generalized additive model. The size of the circles is proportional to the number of excess deaths due to PIC in each state. The color depicts the month March-July) – earlier months are darker colors; later months are lighter colors.

### State Vaccination Guidelines

48 states included criteria as least as inclusive as ACIP guidelines in their first phase of vaccination rollout, which dropped 31 to in the second phase, and 30 in the third phase. *Per capita* dose administration was highest in Alaska in December (9422 doses per 100,000 population); in Washington D.C. in January (13108 per 100,000); in New Mexico February (16579 per 100,000) and lowest in Ohio in December (2490 doses per 100,000 population); Idaho in January (2401 per 100,000); in Arkansas in February (7947 per 100,000; Figure 5). State guidelines at least as inclusive as ACIP were not associated with administered COVID-19 vaccine doses *per capita* each month in correlation tests, nor overall in the negative binomial model. Rollout of the staged, phased vaccine administration prioritization schemes varied. All states are still in Phase 1, but have broken this into three or more stages: as of February 15, 2021, 4 states were on the first stage of rollout, 43 on the second, 4 on the third (Figure 6). 14 states still did not have publicly available, fixed criteria prepared for their third stage of rollout, of 13 which were already actively in the second stage of rollout, however, most of these states have further subdivided their stages into tiers of prioritized groups, and the timeframe for implementing each tier, stage, or phase is largely unknown.

**Figure 5.**
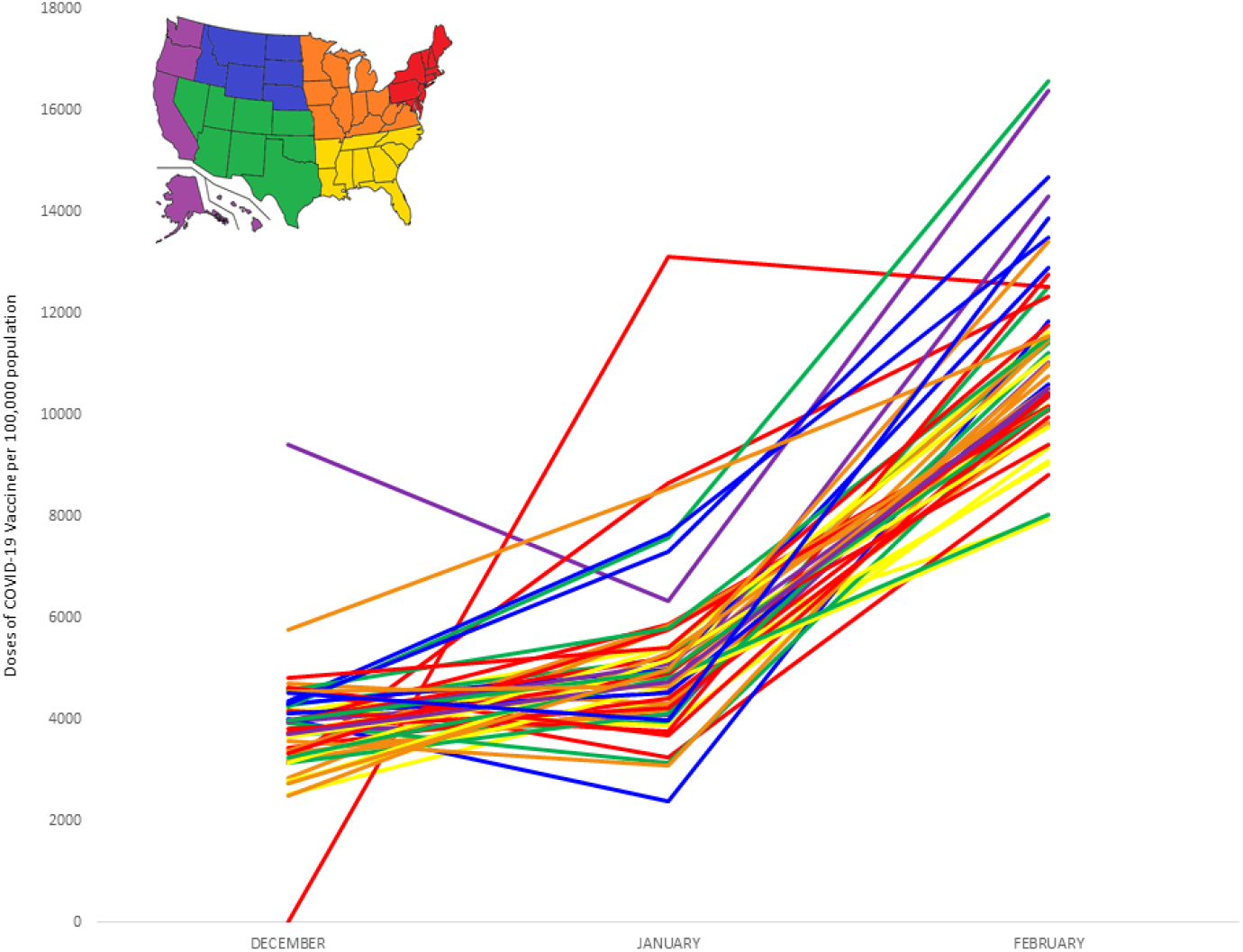
COVID-19 vaccine doses administered per 100,000 population in the United States, December 2020-February 2021. Each state has a line; colors of lines are determined by geographic region per the inserted map.

**Figure 6.**
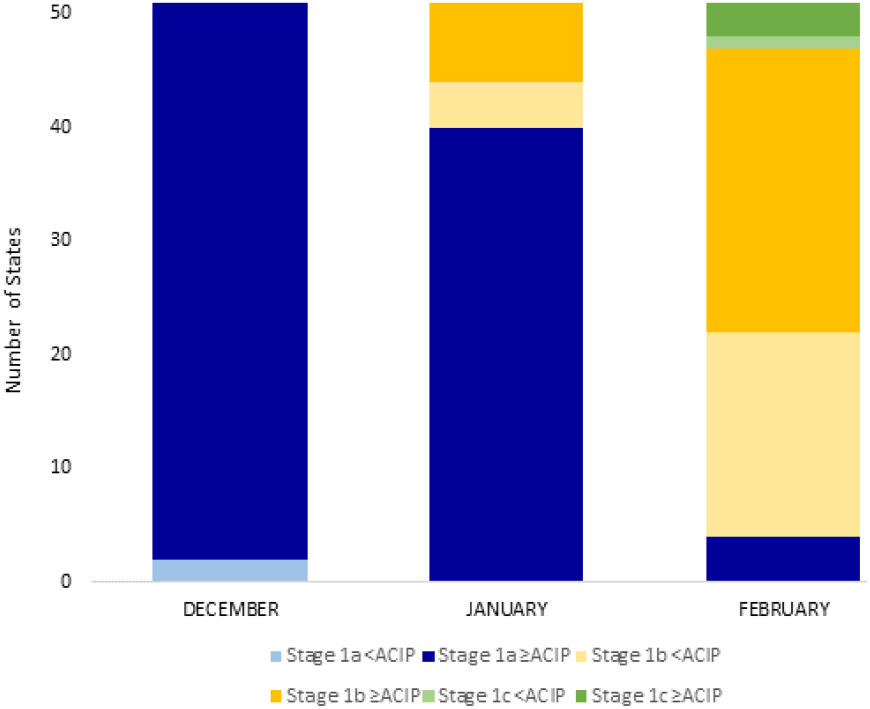
Count of states in each phase of COVID-19 vaccine administration rollout, December 2020-February 2021. Darker colors represent states that were at least as inclusive as ACIP with their vaccine prioritization guidelines; pastel colors represent states that were more restrictive than ACIP guidelines.

The gap between delivered doses (doses received by states) and administered doses (doses given to patients) varied extensively between states in January and February. In January, the median percentage of doses *per capita* that were administered was 59% (minimum: 49% (MO) maximum: 85% (ND)), and by February, the median percentage of total administered doses *per capita* increased to 85% (minimum: 72% (DC) maximum: 98% (NM and ND)). In February by itself, 48 states administered more than the doses that had been delivered during that month (presumably doses that were delivered in previous months were administered in February), showing a ramp-up in vaccine administration.

### Association of per capita vaccination with per capita testing

State-level *per capita* COVID-19 testing was associated with per capita COVID-19 vaccines administered, both overall (rho: 0.71, 95%CI: 0.59-0.81) and for the first month of testing (March 2020) and vaccine (December 2020) rollout (rho: 0.34, 95%CI: 0.03-0.59). The negative binomial model showed that *per capita* testing/epidemic intensity was positively associated with *per capita* COVID-19 vaccine administration in the first three months of rollout (IRR: 1.17, 95%CI 1.10-1.25). States with less inclusive access to COVID-19 testing for the general public (score 0 or 1) were not associated with states with vaccine guidelines less inclusive than ACIP regulations during the first three months of rollout.

## Discussion

In order to obtain accurate statistics on the COVID-19 pandemic, viral testing needs to scale with epidemic intensity. Particularly in the early months of the pandemic, restrictive state-level testing recommendations and difficulties in accessing tests resulted in inadequate testing levels.^5^ This was manifest in higher viral positivity rates and greater differences between reported COVID-19 deaths and excess PIC deaths. From March-May, testing *per capita* and completeness of mortality data both increased, though afterwards this apparent relationship became less clear. It is possible that other characteristics of healthcare systems in each state influenced both the testing rates and completeness of death reporting (i.e. supply chain or personnel issues). Expanded access to testing plays a critical role in obtaining an accurate picture of the progression of the epidemic.

Vaccination prioritization and allocation are, similar to testing recommendations, the responsibility of the states, and have had a similarly unbalanced and inconsistent rollout.^18^ The distinct difference between the strong associations between inclusive testing policy and tests performed *per capita* and the lack of association between inclusive vaccination policy and vaccines administered *per capita* may indicate that other factors, such as deficiencies in the vaccine administration supply chain, lack of access, or vaccine hesitancy may have a stronger role in the variation in vaccines administered *per capita*.

The positive association of testing *per capita* and vaccination *per capita* is of interest, as when both are low, it could be an indication that state-level public health infrastructure is overtaxed, impeding a rapid rollout of these crucial programs. The nature of this impediment could include political, economic, logistic, or human factors, each of which would require a separate course of action to mitigate. Rather than utilizing limited public health resources and personnel to independently construct 50 separate plans and 50 separate prioritization schemes of varying criteria and complexity, it would likely be more efficient if the United States would unite behind a common set of guidelines and standards.^19,20^

This study has certain limitations. Changes in completeness of death reporting could be influenced by other factors that might also vary over time, such as state-level guidelines for coding deaths as due to COVID-19. The viral testing criteria established by state health departments are sometimes superseded by local governments or private clinics.^9^ The testing data counts tests, not individuals, so people tested multiple times will be included multiple times. The vaccine administration data counts total doses administered, and since both of the vaccines currently given Emergency Use Authorization in the United States require two doses, there could be undetected variation in the number of people receiving their first versus their second dose in each state.

Improving COVID-19 vaccine uptake in the face of not only logistical and administrative challenges, but also sociobehavioral issues like hesitancy and misinformation, are further impediments to mass vaccination efforts.^21^ United, clear messaging is essential to build trust and deliver compassionate, competent information to encourage people to get vaccinated. Developing 50 different state-level uptake improvement plans seems like a misallocation of resources. Avoiding this requires that centralized guidance be of high quality and locality-specific adaptability. After a tumultuous pandemic year, resulting in over 500,000 lives lost, it would be unwise to further rely on overextended state-level public health infrastructure to be the ultimate source for policy, implementation, and advocacy.

## Conclusions

The patchwork of state-level testing guidelines was associated with local variations in testing intensity, and these variations had consequences for the completeness of data on key indicators of the severity and progression of the pandemic. By contrast, the uneven state-level vaccination prioritization policies were not associated with variations in *per capita* doses administered. However, vaccination *per capita* correlated with testing *per capita* in the first three months of onset. More coordinated national policies could strengthen the pandemic tracking and response, and prevent us from making the same mistakes again.

## Data Availability

All data and sources are included in this manuscript.

## Authors’ contributions

SP conceived of the study, extracted the data, ran the analyses, and wrote the first draft of the paper. DMW contributed to the design of the study and revisions of the manuscript and plotting.

## Acknowledgements

We would like to thank the state health departments in the United States and the District of Columbia and the COVID Tracking Project for providing data used for these analyses.

## Supplemental Materials

**Supplementary Table 1.**
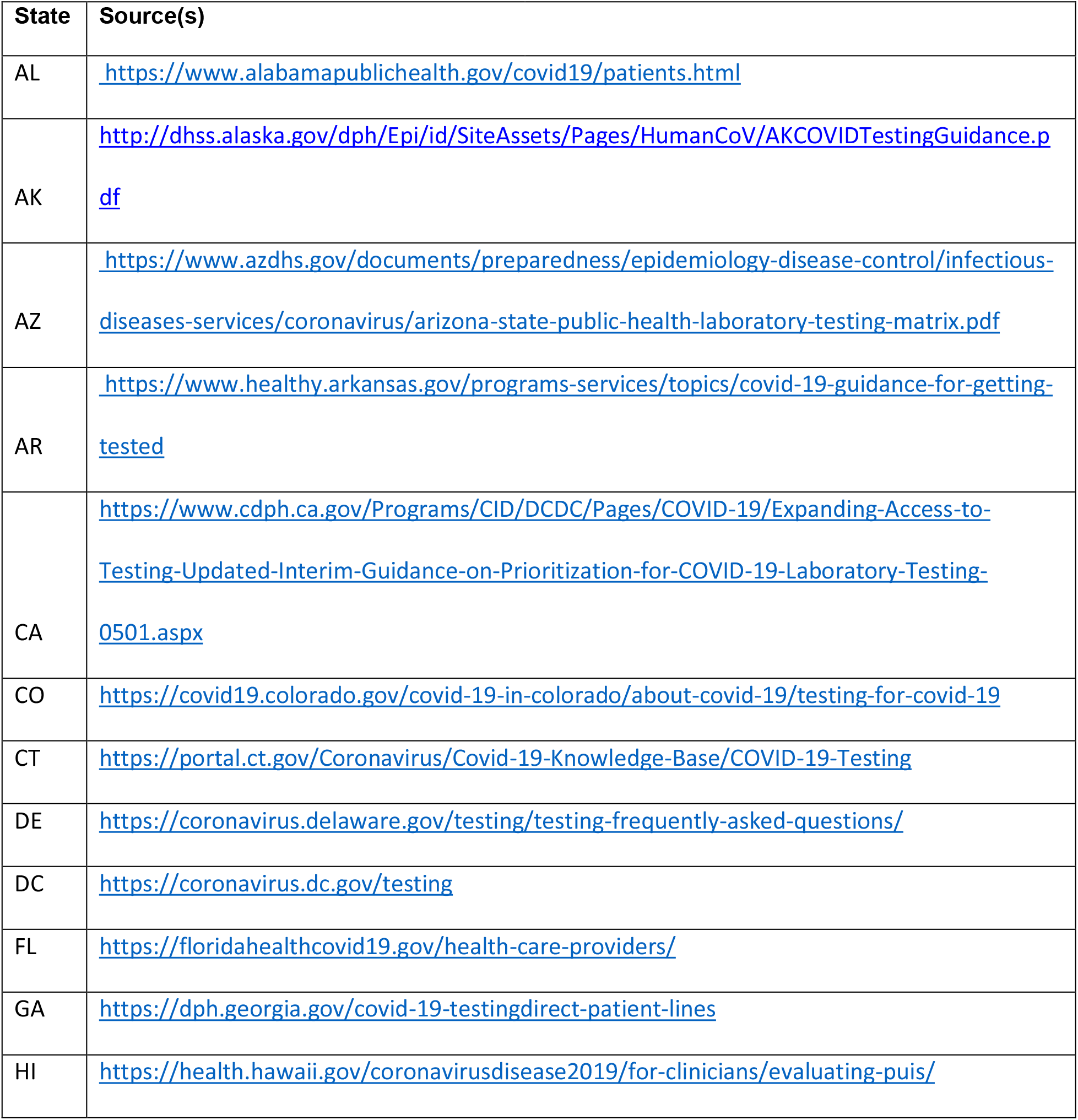
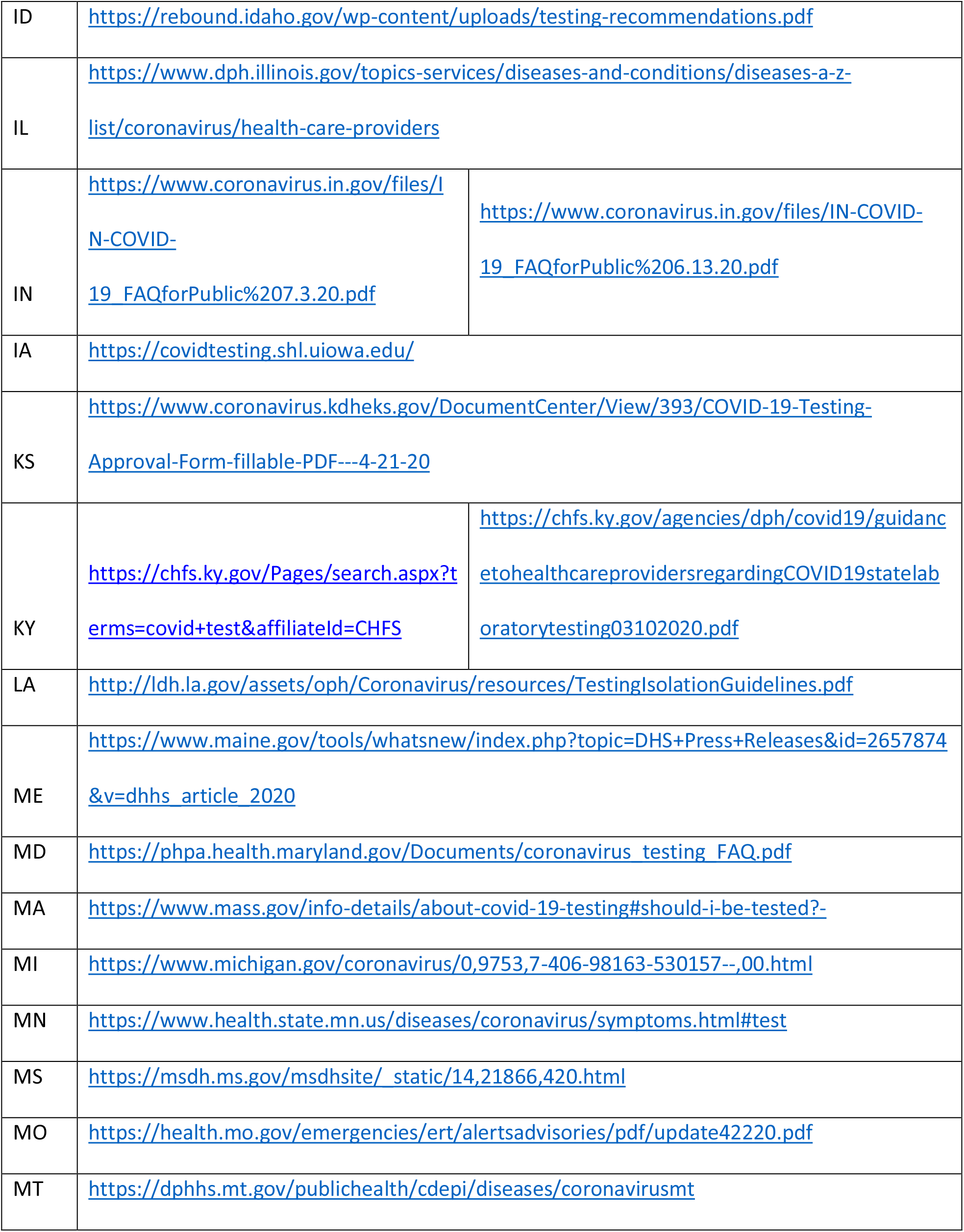
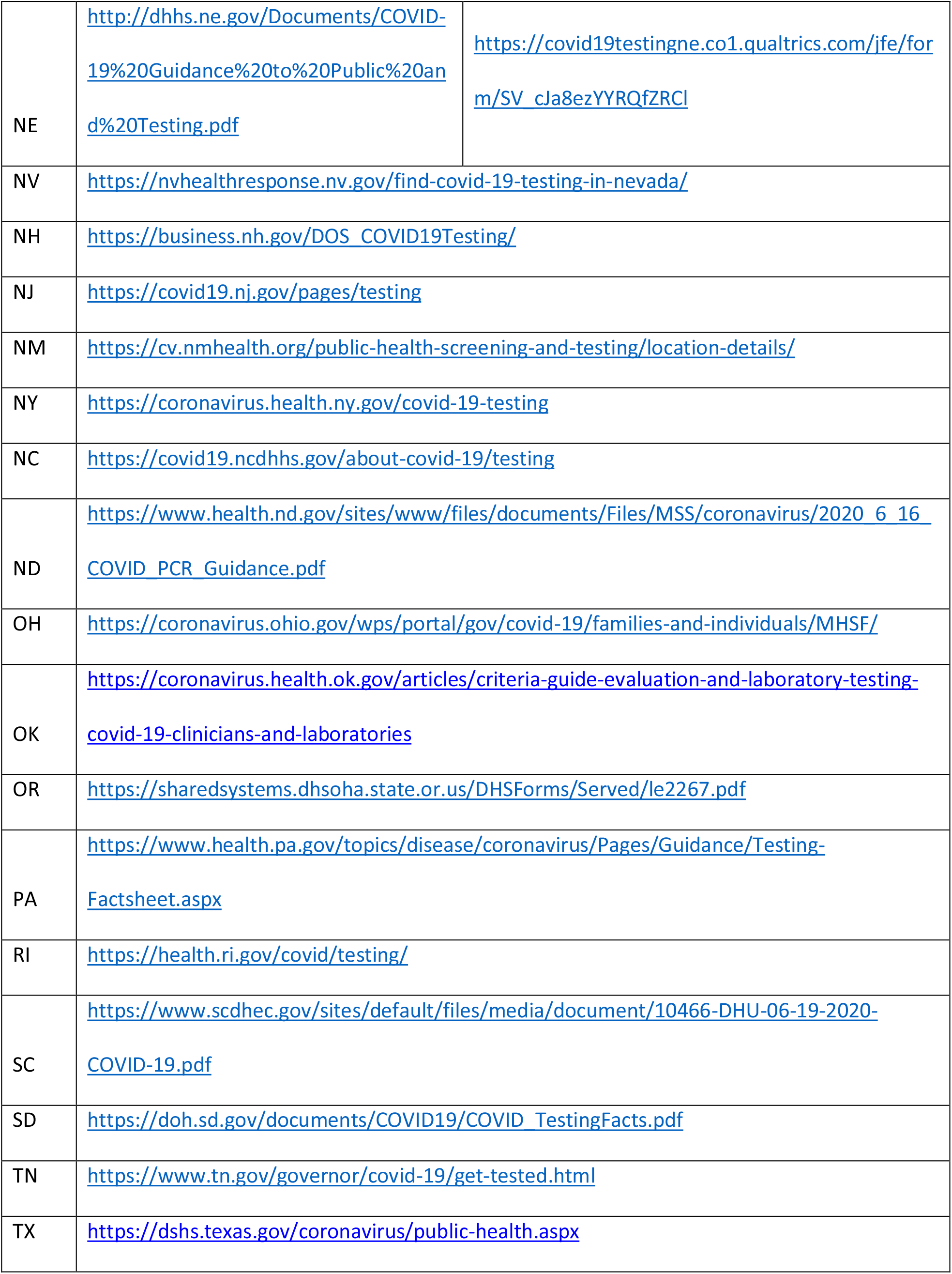
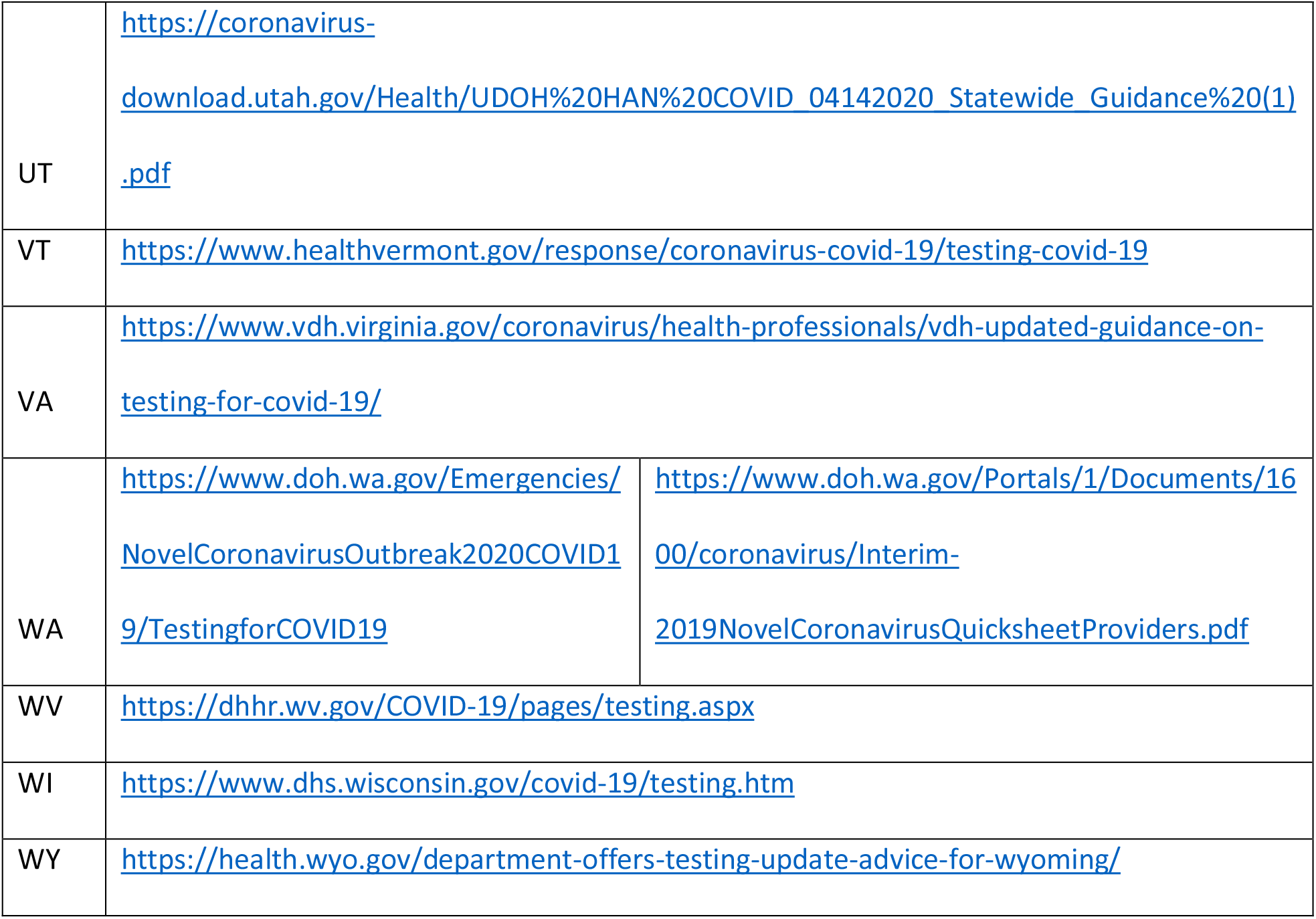
State health department websites, used as the source to determine state-level COVID-19 testing guidelines.

**Supplemental Figure 1.**
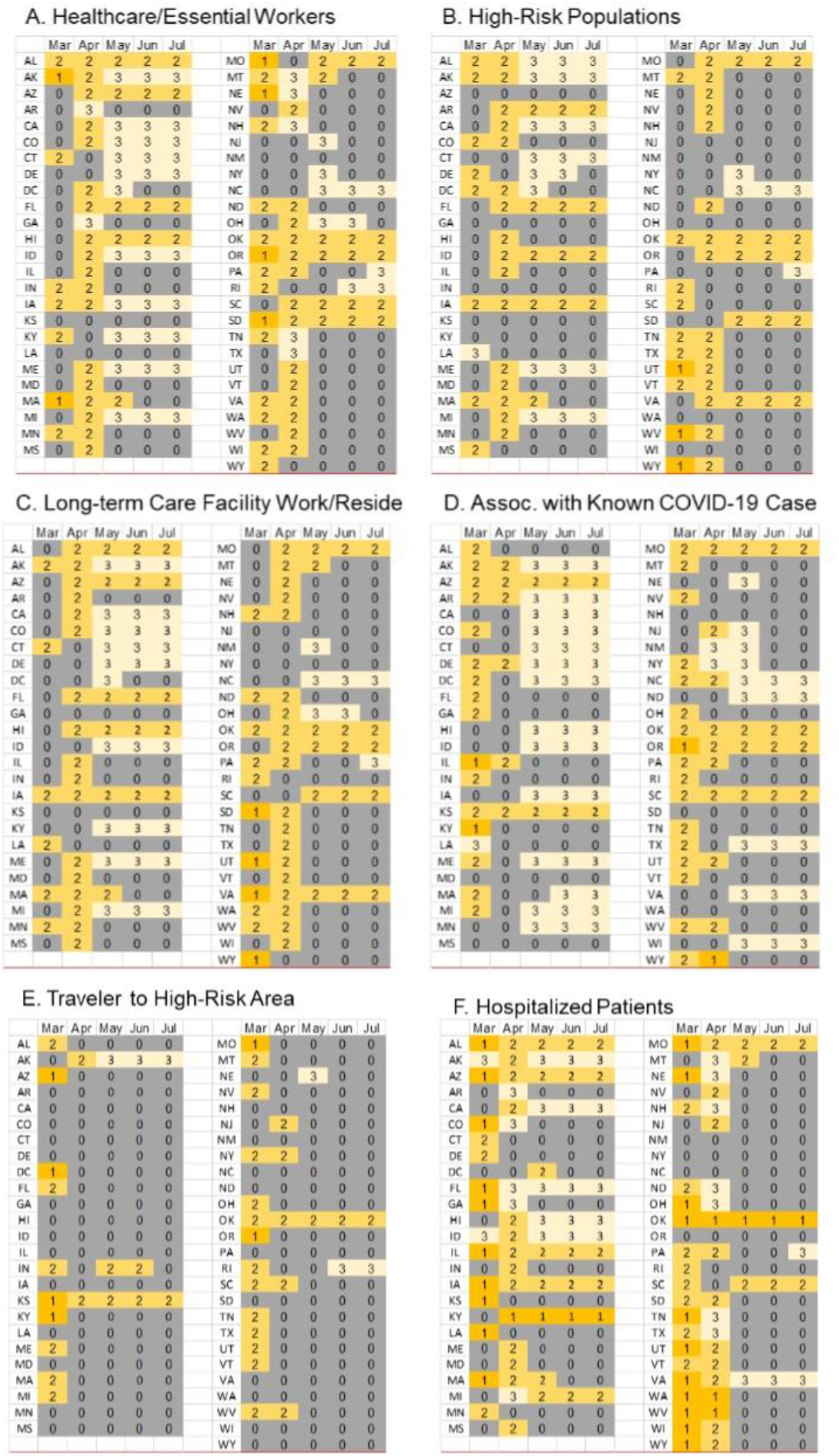
State-level SARS-CoV-2 testing recommendations for selected risk groups, March-July 2020. Criteria for (A) Healthcare/Essential Workers, (B) High-Risk Populations, (C) Long-term Care Facility Residents or Workers (D) Associates of Known COVID-19 case, (E) Travelers to High-Risk Areas, and (F) Hospitalized Patients were scaled as follows: 0 = Criterion not listed, 1 = Negative influenza test required, 2 = Symptomatic individual, 3= Any individual.

